# Adaptation and psychometric evaluation of the ENhancing Assessment of Common Therapeutic factors (ENACT) tool to build the capacity of primary care physicians in low-resource settings

**DOI:** 10.64898/2026.01.20.26344430

**Authors:** Asma Humayun, Asma Nisa, Israr ul Haq, Arooj Najmussaqib, Noor ul Ain Muneeb

## Abstract

**Background:** With the global focus on task-sharing initiatives to bridge the treatment gap, the assessment of competencies of non-specialists is essential. The ENhancing Assessment of Common Therapeutic factors (ENACT) was developed to assess the competence of non-specialists to deliver psychosocial interventions, but the evidence regarding its application, implementation and adaptation process is limited. The present study aims to fill this gap by adapting and pilot testing the ENACT tool for resource-constrained settings.

**Methods and Results:** We used a sequential, mixed-methods approach to adapt and pilot test the ENACT tool for evaluating therapeutic skills of non-specialists trained in mhGAP trainings. The adaptation process consisted of selection of competencies through literature review and focus group discussion. This was followed by designing a test strategy comprising development of a case vignette, its iterative refinement and adapting scoring framework. Lastly, feasibility was tested in the mhGAP workshop setting.

Eight competencies were retained based on their relevance, comprehensibility and feasibility. Each competency was rated on a four-point ordinal scale, with level 1 indicating no skill, while 2, 3 and 4 reflecting some, all and advanced skills, respectively. Role-plays were video-recorded and rated by trained raters. Good inter-rater reliability and significant correlation between competencies were observed.

**Discussion/Conclusion:** ENACT can be used to assess therapeutic skills but must be paired with structured rater training and contextual adaptation. Our study addressed its implementation challenges by making it manageable with limited resources, time and raters, making it scalable for low-resource settings. Our findings can inform mhGAP training design and delivery, facilitate in shifting its emphasis from a biomedical centered training to patient-centered approach. It can also guide curriculum refinement, identify priority skill areas for refresher trainings, and serve as mechanism for quality improvement for training and supervision. Lastly, it can inform policy decisions to integrate foundational competencies into large-scale training and supervision systems.

## INTRODUCTION

To address the massive treatment gap of mental disorders in low-and-middle-income countries (LMICs), World Health Organization (WHO) recommends integration of mental health into primary care. To achieve this, building the capacity of primary care physicians (PCPs) to identify and manage mental health conditions is crucial since they are the first point of contact for many people with mental health conditions in LMICs (World Health Organization, 2021a).

The ENhancing Assessment of Common Therapeutic factors (ENACT) tool has been widely used to assess competence of non-specialists for task-sharing initiatives to deliver psychological and psychosocial interventions in low-resource and humanitarian settings (Jordans et al., 2022; Kohrt, Jordans, et al., 2015; Pedersen et al., 2021; Spedding et al., 2022). Skill assessment of non-specialist prior to training can guide focus and duration of training, supervision, and the selection criteria of non-specialists to improve their competence in task-sharing (Rose et al., 2023). The World Health Organization (2017) also recommends peer- and-facilitator assessed role-plays to help develop skills of PCPs for implementing mhGAP guidelines. However, most of mhGAP-related initiatives in different settings have relied on self-reported measures to assess knowledge and attitudes of non-specialists (Keynejad et al., 2018). Kohrt and colleagues (2018) addressed this gap by assessing competency of non-specialists using ENACT for mhGAP trainings in Uganda, Liberia and Nepal. They used contextualized and standardized role plays to apply ENACT but did not document the process of adapting the protocol in their study.

Others have highlighted that using ENACT without contextual modification is challenging when applied to specific training models (Pedersen et al., 2021; Spedding et al., 2022). For instance, some competencies may not align optimally with the scope of mhGAP training for non-specialists or may be difficult to operationalize within brief standardized role-play assessments. Furthermore, previous studies have also highlighted the need to adapt scoring structures and administration procedures to enhance feasibility, rater usability, and discriminative capacity in diverse implementation settings (Kohrt et al., 2025; Spedding et al., 2022). Several considerations, such as quality improvement for training and supervision; scalability and user-friendliness by limiting the number of items; using a scoring system that captures skills hierarchies; and minimizing subjective appraisals, have been highlighted for the adaptation of ENACT (Kohrt, Ramaiya, et al., 2015). These considerations underscored the need for a systematic adaptation and psychometric evaluation of the ENACT tool.

The present study aims to adapt and pilot test the ENACT tool for evaluating ENACT competencies, (here onwards termed as foundational therapeutic skills), particularly when delivered by non-specialists, as part of implementation of mhGAP-HIG guidelines in resource-limited settings facing humanitarian challenges.

## METHODS

We used a sequential, mixed-methods approach to adapt and pilot test the ENACT tool for evaluating foundational therapeutic skills of PCPs trained in the mhGAP Humanitarian Intervention Guide (mhGAP-HIG) in Pakistan. The study is described in two parts: the process and outcome of systematic adaptation of the ENACT tool and pilot testing it within mhGAP-HIG training setting. The pre and post assessment of ENACT in mhGAP context is published elsewhere.

Our objective of the adaptation process was not to retain all therapeutic competencies included in the original ENACT (Kohrt, Jordans, et al., 2015), but to identify a set of foundational competencies that could be reliably demonstrated and tested within mhGAP-HIG training contexts. Given the biomedical orientation of primary care practice (Nawaz et al., 2020) and the brief, standardized nature of role-play assessment, we prioritized feasibility, observability, and contextual relevance over exhaustive coverage of competencies.

### Part I: Adaptation of the ENACT tool

We followed three steps to adapt the ENACT tool to ensure its contextual relevance, feasibility, and suitability for assessing key communication and therapeutic competencies among PCPs in low-resource settings (Figure 1):

**Figure 1:**
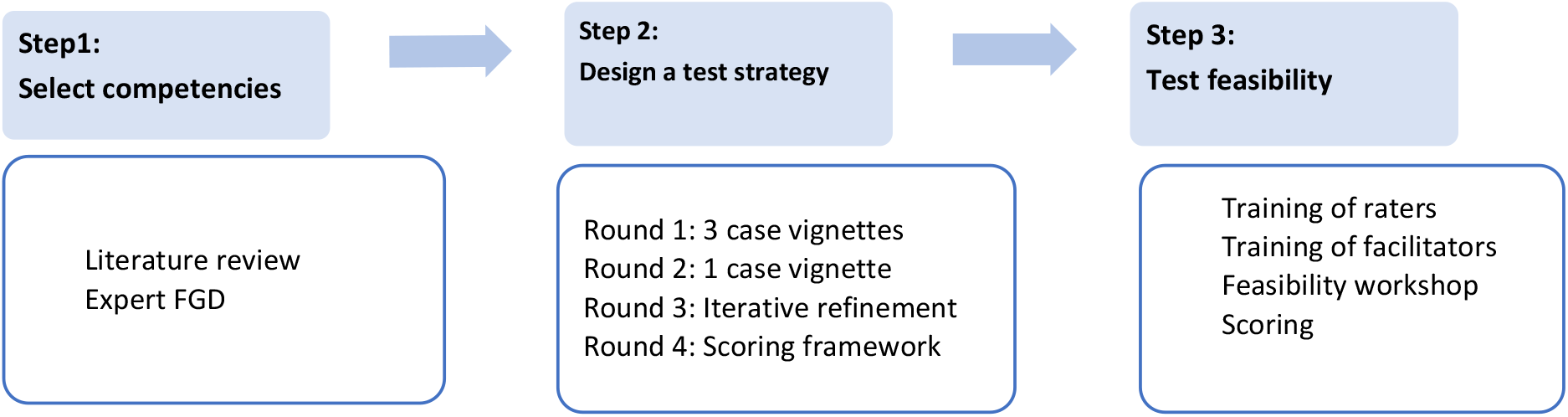
Process of adaptation

Step 1: Selection of competencies

Step 2: Designing a test strategy

Step 3: Feasibility testing

We followed the approach used for adaptation of ENACT to develop EQUIP through developing consensus, selecting competencies, and evaluating their feasibility using role plays (Kohrt et al., 2025). Data from all steps of the adaptation and feasibility process were analyzed using descriptive methods. No inferential statistical analyses were conducted during the adaptation stages.

Findings from these descriptive methods were used to refine competency selection, vignette design, scoring procedures, and administration protocols, and to confirm readiness for subsequent pilot testing with a larger sample.

#### Step 1: Select competencies

The core team (IH, AN, NM) reviewed prior ENACT studies to identify competencies that had already been documented as redundant or overlapping and were therefore potential candidates for exclusion or modification and identified three competencies from the original 18-item ENACT for provisional exclusion. These included use of praise or positive reinforcement, managing the ending of the session, and structured session management behaviors (e.g., agenda-setting). They agreed with prior ENACT adaptations that these competencies may be redundant with broader relational competencies, difficult to observe reliably within brief standardized role-play assessments, or conceptually overlapping with other relational or problem-focused competencies, particularly in task-sharing and primary care contexts (Kohrt et al., 2015; Kohrt et al., 2015b; Spedding et al., 2022).

Then we conducted a review with three experts including a psychiatrist (AH), a clinical psychologist and a researcher (AsN+ AN), each with a minimum of five years’ experience in mental health care. Two experts had mhGAP training experience with PCPs as well, while the researcher had expertise in cultural adaptation of assessment tools.

The experts independently rated each of the 15 competencies on a four-point scale (1 = not relevant/clear/feasible, 2 = somewhat relevant, 3 = quite relevant, 4=highly relevant) against a pre-defined criterion: (1) Relevance, defined as alignment with the objectives of the mhGAP-HIG implementation in our healthcare and cultural context; (2) Clarity and comprehensibility of integrating these competencies into a standardized role-play scenario; and (3) Feasibility, defined as the practicality of testing a competency within a resource-limited role play assessment without compromising the evaluation of other competencies.

During expert review, competencies receiving consistent high ratings (high content validity index score of 3 or 4) across relevance, comprehension, and contextual suitability were retained, while those with consistent lower ratings (score of 1) were excluded. A subset of competencies received moderate ratings on one or more criteria. For these competencies, experts provided the rationale for rating the competencies, as needed. These competencies were discussed by the research team to develop consensus for lower ratings, such as contextual incongruence, or challenges in comprehension or application or overlapping reasons. For example, the competency “assessment of harm to self, harm to others, and from others, and development of a collaborative response plan” was considered too complex to be demonstrated through an integrated role-play. Similarly, “appropriate involvement of family members and other close persons” was not deemed feasible in a role play because of time constraints, and not because it was not relevant or understandable. Competencies receiving moderate ratings were considered important clinically, but excluded because these required higher-order cognitive integration, longitudinal engagement, or system-level interaction that could not be meaningfully demonstrated within a brief, single-encounter role-play. Based on ratings by the experts, we retained eight competencies as the final set to be operationalized in this study. Figure 2 shows the process of selection of these competencies.

**Figure 2:**
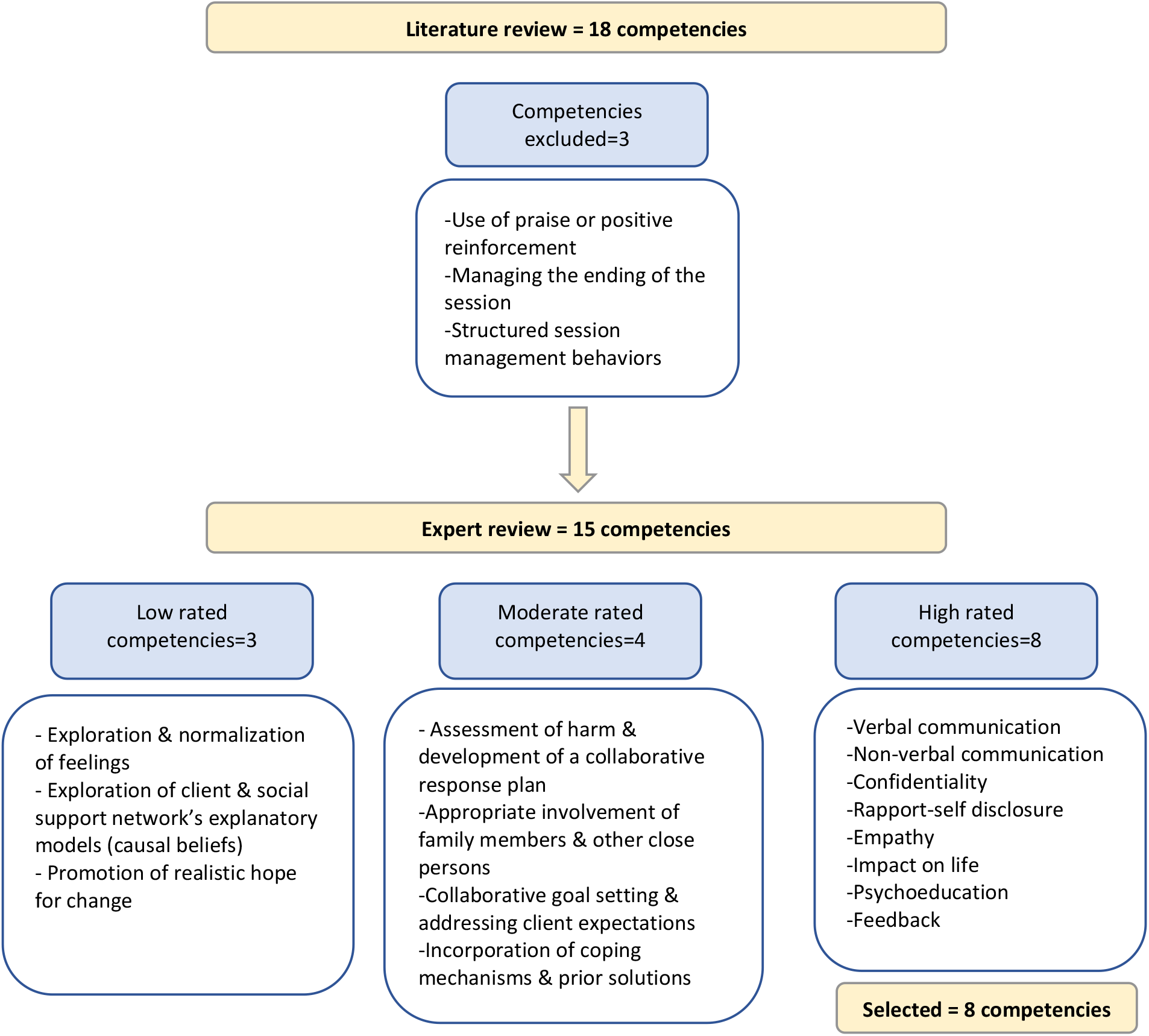
Select competencies

#### Step 2: Design a test strategy

In the first round, the core team developed three separate case vignettes to integrate the 8 competencies. Internal role-play testing indicated that administering these vignettes was resource and time-intensive and checked overlapping skills across scenarios.

##### User-centered design of the vignette

In the second round, we developed a single case vignette format to demonstrate all eight retained competencies within one role play of a clinical encounter. We employed user-centered design, as advised by Moore at al., (2015). Our aim was to avoid spending role play time on diagnosis, and focus on demonstrating key foundational skills, so we developed a structured vignette, as suggested by Kohrt et al. (2018), around a case of epilepsy, a condition commonly managed by PCPs in our healthcare context. It was designed in Urdu, aligned with routine communication practices of PCPs in Pakistan. The vignette-based role-play functioned as an assessment tool to standardize clinical input, reduce variability related to case complexity, and isolate interactional competencies from diagnostic reasoning. By controlling the clinical scenario, the assessment focused on the skills rather than clinical decisions.

##### Standardization of the vignette

In the third round, the vignette underwent revisions to reduce unnecessary clinical detail, refine prompts, and standardize flow. We incorporated standardized actor prompts to minimize variability across facilitators and provide similar opportunities to all participants, as recommended (Kohrt et al., 2025). This approach aligns with ENACT’s emphasis on observable behaviors and supports inter-rater reliability by reducing context-driven variability across assessments. Iterative role-plays conducted by the core team were used to examine clarity of prompts, reduce redundancy, and optimize the administration time to approximately5 minutes per role-play (Evans et al., 2015). The process of iterative refinement was followed by a final expert review.

##### Scoring framework

The core team also reviewed the literature to examine the suitability of the original ENACT scoring framework for brief, standardized role-play assessments in mhGAP-HIG training settings. This review drew on the original ENACT development and scoring papers, subsequent ENACT adaptations, and EQUIP-related competency assessment work that documented challenges in applying the original response options in practice (Kohrt, Jordans, et al., 2015; Kohrt, Ramaiya, et al., 2015; Pedersen et al., 2021; Spedding et al., 2022). As a result, the original scoring framework was adapted retaining a four-level structure but redefined Level 1 to indicate absence of the skill or failure to elicit the competency rather than active harm. Levels 2–4 reflected increasing quality of skill demonstration, ranging from partial or basic demonstration to consistent demonstration of all basic skills and inclusion of advanced or nuanced skills. The experts also reviewed the adapted four-level scoring framework in the light of its clarity, usability, and suitability to rate a range of competency.

#### Step 3: Test feasibility

After developing a strategy to administer the adapted ENACT tool, we conducted a small-scale feasibility assessment to evaluate the procedure and address practical challenges for implementation.

To ensure a standardized administration of the ENACT role-play vignette and consistent evaluation of participants, scoring procedures, we held a two-day virtual training to orient the raters to the adapted ENACT tool and scoring framework. The raters included three mental health professionals, each with a minimum of five years’ experience in mental health practice and research. Raters were provided with ENACT training materials, including sample role-play videos and supporting resources, to facilitate familiarization with observable competency indicators and scoring anchors(World Health Organization, 2021b). Raters then participated in structured practice sessions, working in groups of three (an actor, a PCP, and a rater) to apply the scoring framework under simulated conditions. Following each practice session, we reviewed ratings and addressed discrepancies to develop a shared understanding of scoring criteria.

##### Formative evaluation

A feasibility workshop was conducted on 20 PCPs selected through purposive sampling. They included 12 males and 8 female physicians with a mean age of 37.35 ± 10.28 years and varying levels of experience: 9 participants with <5 years; 6 with 5-10 years; and 5 with >10 years of practice. The single-vignette role-play was administered by two mhGAP facilitators at two separate stations in a training-like setting. As recommended, all role-plays were video recorded with verbal consent from participants for adaptation and research purposes (Kohrt, Ramaiya, et al., 2015).

The assessment was administered by two mhGAP facilitators (as actors), who were able to assess approximately 10 participants within one hour, including a short break after half of the participants had completed the assessment. The facilitators documented their observations about participant engagement, comprehension gaps in role-play instructions, and administration time per participant.

For the feasibility testing phase, feasibility indicators were summarized descriptively. Administration time per participant and total time required for group administration were recorded using a stopwatch and reviewed against predefined feasibility thresholds. Observations related to participant comprehension of instructions, engagement with the role-play, and completion of the assessment were documented to identify procedural challenges requiring refinement.

All role-plays were independently reviewed by trained raters who applied the adapted scoring framework to assess competencies. The scores by all three raters for each role-play were compared descriptively. This review focused on confirming consistency in scores across competencies and identify any systematic discrepancies or ambiguities in the scoring.

Overall, findings from the feasibility testing supported the procedural practicality of the adapted ENACT administration taking into account the resources, participant engagement, vignette design, and scoring procedures.

### Part 2: Pilot Testing

The objective of the pilot testing phase was to evaluate the psychometric performance of the adapted ENACT tool when administered at scale within mhGAP-HIG training settings, using baseline (pre-training) competency assessments among PCPs.

We employed a purposive sampling strategy to recruit a total of 118 PCPs for pilot testing of the adapted ENACT tool. Baseline (pre-training) competency assessments were collected across seven mhGAP-HIG training workshops conducted in different humanitarian settings across Pakistan.

Participation in the study was voluntary and the purpose and procedures of the study was clearly explained. Written informed consent was obtained for participation and video recording of role-play assessments. All recordings were used solely for research and training evaluation purposes.

We received nominations for PCPs from the respective health departments based on predefined inclusion criteria. Eligible participants were required to hold a recognized professional qualification, demonstrate interest in professional development, show inclination toward a biopsychosocial approach to care, and adhere to ethical standards.

**Table 1.**
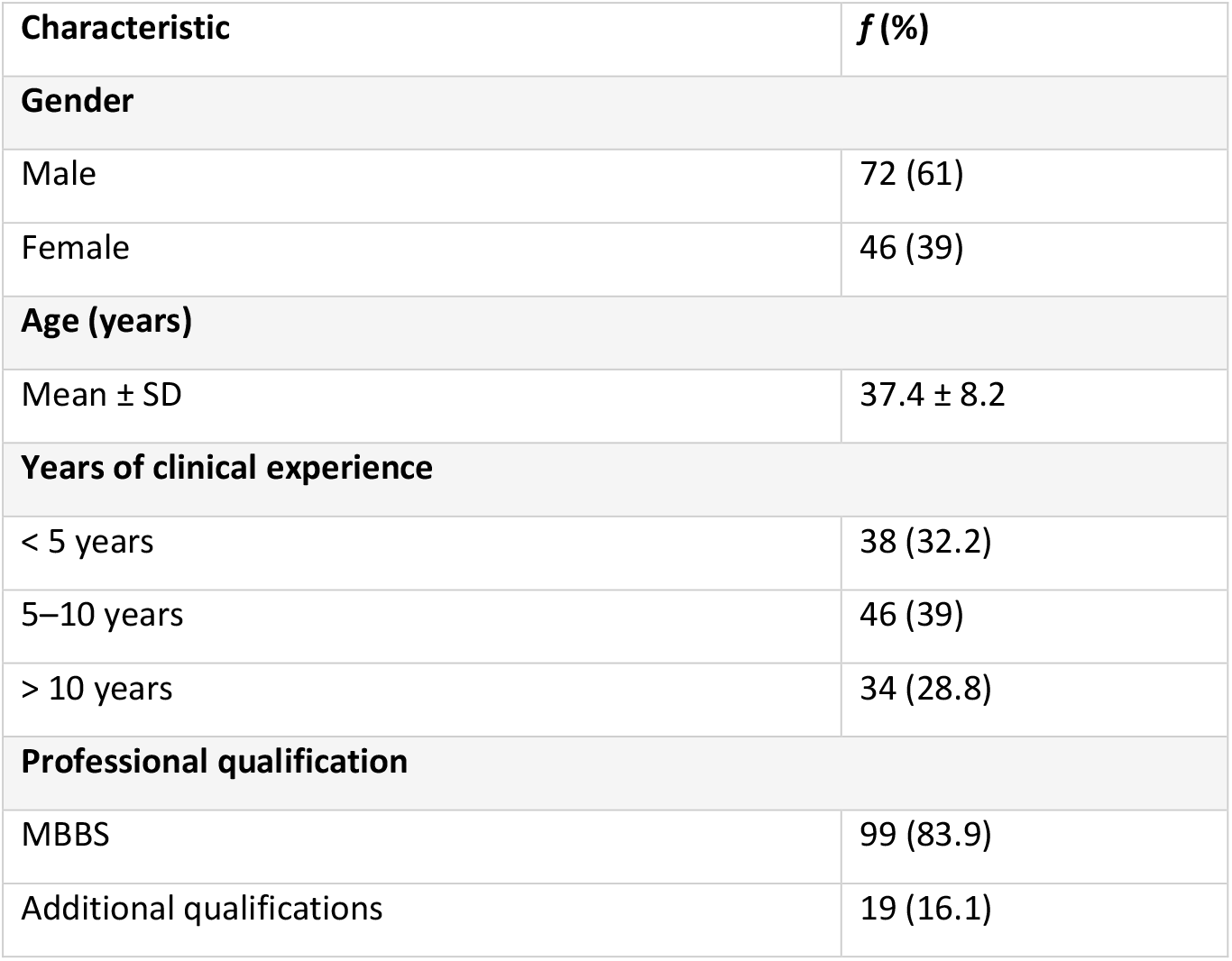
Demographic characteristics of primary care physicians included in pilot testing (N = 118)

| Characteristic | *f* (%) |
| --- | --- |
| **Gender** |  |
| Male | 72 (61) |
| Female | 46 (39) |
| **Age (years)** |  |
| Mean ± SD | 37.4 ± 8.2 |
| **Years of clinical experience** |  |
| < 5 years | 38 (32.2) |
| 5–10 years | 46 (39) |
| > 10 years | 34 (28.8) |
| **Professional qualification** |  |
| MBBS | 99 (83.9) |
| Additional qualifications | 19 (16.1) |

We used the adapted ENACT tool to assess therapeutic competencies, using a standardized, single-scenario role-play vignette administered in Urdu. Each competency was rated using a four-point ordinal scale, where Level 1 indicated absence of the skill or failure to elicit the competency, Level 2 reflected partial or basic demonstration of the skill, Level 3 indicated consistent demonstration of all basic skills, and Level 4 represented comprehensive demonstration including advanced or nuanced skills.

Role-play actors were drawn from a pool of trained mhGAP facilitators who were not involved in scoring to reduce variability across assessments and minimize assessor bias. A total of three actors rotated across sessions to manage workload and maintain consistency. Each assessment was completed within approximately five minutes per participant. Brief breaks were scheduled after every 10 assessments. Role-plays were video recorded to support scoring using the adapted ENACT framework.

The video-recorded role-plays were independently scored by four trained raters using the adapted four-level ENACT scoring framework.

Data analysis focused on evaluating inter-rater reliability, baseline competency performance, and associations among competencies assessed using the adapted ENACT tool. Inter-rater reliability was examined using intraclass correlation coefficients (ICCs) calculated for individual competencies and for the total ENACT score. A two-way random-effects model with absolute agreement was used to assess the consistency of ratings across raters. Baseline competency performance was examined descriptively by calculating means, standard deviations, and score distributions for each competency and for the overall ENACT score, to identify patterns of pre-service skill strengths and gaps among primary care physicians. Lastly, Pearson correlation analyses were conducted to examine associations among competency scores, providing preliminary insight into relationships between different skill domains assessed by the adapted ENACT tool.

**Table 2:** Inter-rater reliability (Intraclass Correlation Coefficients) for ENACT indicators, pre- and post-training (n = 4 raters)

| ENACT Indicator | ICC | 95% CI |
| --- | --- | --- |
| Non-verbal communication | 0.82 | 0.76 - 0.87 |
| Verbal communication | 0.77 | 0.69 - 0.84 |
| Confidentiality | 0.87 | 0.82 - 0.91 |
| Rapport–self disclosure | 0.85 | 0.79 - 0.89 |
| Empathy | 0.82 | 0.76 - 0.87 |
| Impact on life | 0.75 | 0.65 - 0.82 |
| Psychoeducation with local terms | 0.80 | 0.73 - 0.86 |
| Feedback | 0.75 | 0.66 - 0.82 |

Across ENACT indicators, ICC ranged from 0.75 to 0.87, indicating overall good inter-rater reliability among the four raters. Reliability was highest for confidentiality and rapport–self disclosure, while impact on life and feedback demonstrated comparatively lower, though still acceptable agreement.

**Table 3:** Baseline (Pre-training) ENACT competency performance among primary care physicians (N = 118)

| Competency | Mean (SD) | Level 1 n (%) | Level 2 n (%) | Level 3 n (%) |
| --- | --- | --- | --- | --- |
| Non-verbal communication | 1.75 (0.54) | 35 (29.9) | 76 (65.0) | 6 (5.1) |
| Verbal communication | 1.88 (0.44) | 19 (16.2) | 93 (79.5) | 5 (4.3) |
| Confidentiality | 1.62 (0.60) | 52 (44.4) | 58 (49.6) | 7 (6.0) |
| Rapport & self-disclosure | 1.72 (0.47) | 34 (29.1) | 82 (70.1) | 1 (0.9) |
| Empathy | 1.63 (0.57) | 48 (41.0) | 64 (54.7) | 5 (4.3) |
| Impact on life | 1.73 (0.52) | 36 (30.8) | 77 (65.8) | 4 (3.4) |
| Psychoeducation (local terms) | 1.91 (0.44) | 17 (14.5) | 94 (80.3) | 6 (5.1) |
| Feedback | 1.54 (0.55) | 57 (48.7) | 57 (48.7) | 3 (2.6) |

**Note.** Only Levels 1–3 were observed at baseline; no Level 4 demonstrations were recorded.

Baseline ENACT scores indicated variation in competency performance across domains. Communication-related competencies, particularly verbal communication and psychoeducation delivered using local terms, demonstrated relatively higher baseline performance. In contrast, competencies related to feedback, confidentiality, and empathy showed lower baseline performance and were less consistently demonstrated during role-play assessments. Overall, most competencies were demonstrated at partial or basic levels, with limited evidence of consistent skill demonstration across participants at baseline.

**Table 4:** Pearson correlations among baseline ENACT competencies (N = 118)

| Competency | 1 | 2 | 3 | 4 | 5 | 6 | 7 | 8 |
| --- | --- | --- | --- | --- | --- | --- | --- | --- |
| 1. Non-verbal communication | — |  |  |  |  |  |  |  |
| 2. Verbal communication | .02 | — |  |  |  |  |  |  |
| 3. Confidentiality | .29** | -.01 | — |  |  |  |  |  |
| 4. Rapport–self disclosure | .16 | .04 | .32** | — |  |  |  |  |
| 5. Empathy | .38** | .03 | .37** | .19* | — |  |  |  |
| 6. Impact on life | .09 | .04 | .19* | .35** | .24** | — |  |  |
| 7. Psychoeducation (local terms) | .38** | .03 | .22* | .33** | .21* | .19* | — |  |
| 8. Feedback | .25** | .09 | .35** | .39** | .28** | .28** | .36** | — |

**Note.** $p < .05$ , $p < .01$ (two-tailed).

Correlation analyses indicated that several competencies demonstrated moderate, statistically significant associations at baseline. Competencies related to confidentiality, rapport-self disclosure, empathy, assessment of impact on daily life, psychoeducation using local terms, and feedback were positively correlated, suggesting that these skills tended to co-occur during role-play assessments. In contrast, verbal communication showed weak and non-significant associations with most other competencies. Non-verbal communication demonstrated moderate associations with multiple competencies, indicating overlap with broader relational skill demonstration.

## DISCUSSION

ENACT is designed to assess observable competencies for non-specialists in task-sharing initiatives, but requires pairing with strong rater training and contextual adaptation (Kohrt, Jordans, et al., 2015; Mwenge et al., 2022; Spedding et al., 2022). A key challenge in scaling up psychological interventions is the evaluation of the competencies among newly trained workforce engaged in service delivery (Ottman et al., 2020). This study contributes a pragmatic, implementation-oriented model for adapting ENACT that balances psychometric robustness with feasibility within mhGAP-HIG training contexts, in resource-limited settings.

The use of ENACT is resource and time-intense, requiring supervisors and raters for its implementation, burdening the training in low-resource settings for administration (Adewuya et al., 2025; Rahman et al., 2019), hindering in its scalability. The evidence highlights the need and adaptation process, but fails to documents steps to replicate or implement in low-resource settings or for mhGAP context (Kohrt, Mutamba, et al., 2018). Our study offers a replicable model for making competency-based assessment operationally realistic and scalable across diverse low-resource settings, strengthening the quality of integrated mental healthcare.

Despite the emphasis on strengthening and measuring knowledge, attitude and skills for effective implementation of mhGAP guidelines (World Health Organization, 2017), there are limited approaches to assess newly acquired skills in routine practice, particularly in resource-constrained settings. The outcome of mhGAP implementation is frequently limited to knowledge gains, with comparatively less evidence on actual competency enactment in clinical encounters, underscoring the need for practical and scalable assessment methods (Keynejad et al., 2018; Petagna et al., 2023). Mere reliance on self-reported measures undermines the translation of training into practice (Kohrt, Mutamba, et al., 2018; Le et al., 2022). Role-plays have the benefits of standardization and assessment of observed behavior rather than written knowledge but are a resource intense strategy (Ottman et al., 2020). The routine application of competency-based tools in evaluating the performance of non-specialists is often constrained by time, human resource, and supervisory requirements needed for administration (Adewuya et al., 2025; Le et al., 2022).

As recommended by Kohrt and colleagues (2018), we have contextualized the ENACT competencies considering routine primary care workflow, and embedded it in a role-play assessment as a feasible and resource-efficient assessment strategy within mhGAP training contexts, overcoming additional resource, supervisory or time requirements. This approach aligns with key implementation principles by being feasible, acceptable, and relevant to routine practice. We also considered alternative formats, such as individual task-based demonstrations of specific competencies; however, were neither feasible nor practical within mhGAP-HIG training settings

Our adaptation process comprised of adapting the existing items (Asher et al., 2021; Kohrt et al., 2020) rather than starting from scratch (Spedding et al., 2022). To establish the psychometric performance of the scale in our context, inter-rater reliability and content validity was established using both quantitative and qualitative approach (Haynes et al., 1995). We agreed with Laurenzi et al.(2025) that a shorter, relevant, feasible and pragmatic version is more helpful to our context. Using a vignette in local language was a critical component of our adaptation process as it enhances cultural and linguistic relevance of the assessment (Kohrt, Jordans, et al., 2015). As recommended by Kohrt et al. (2025), the prompts were flexible and adaptable, in the language typically used by service users, while the items that were not feasible to assess were dropped from the tool. It also reduced cognitive burden, facilitated natural communication, and improved ecological validity of observed behaviors.

In addition, we coded the competencies directly from video recordings rather than following a translated or transcript-based approach. Unlike other studies who used audio recordings (Laurenzi et al., 2025; Mwenge et al., 2022), we video recorded the sessions to help assess non-verbal skills and relational features such as warmth, responsiveness, and empathic engagement.

We opted for a revised scoring framework as our findings were consistent with other studies (Kohrt, Ramaiya, et al., 2015; Mwenge et al., 2022) about the lowest rating category (of harmful behaviour) that are rare in training settings, likely to make raters uncomfortable; and the use of this category is constrained by social desirability and cultural norms in training contexts. Difficulties in distinguishing between absence of a competency and demonstration of harmful behavior in brief standardized role-plays has also been reported (Kohrt, Ramaiya, et al., 2015; Pedersen et al., 2021). As recommended by Kohrt et al. (2015), user-friendliness, limited items and scoring system that reflects hierarchy of skill was developed. The original scoring did not show any difference in no skill or some basic skills, both being scored as level 2 (World Health Organization, 2021b). The adapted scoring facilitates optimum skill measurement, as well as precisely captures the difference in pre and post training scores, suited for mhGAP training, where competency assessments were to be conducted at predefined time points. The revised scoring framework was designed for formative training evaluation rather than monitoring harmful practices (Jordans et al., 2022).

As a prerequisite for sustained use of competency-based assessments, feasibility was hallmark of this adaptation (Jordans et al., 2022; Peters et al., 2014). We relied on recordings to reduce supervisory and logistical burden, administering on a large sample without compromising the training time, making it scalable for mhGAP programmes (Laurenzi et al., 2025; Mwenge et al., 2022). We limited the administration time to 5 minutes per participant, even shorter than Kohrt et al., (2018), and created multiple stations to apply a theoretically robust tool operationally practical. Though minor hesitation was observed but neither the trainers nor the participants found the execution or scoring of the role-plays difficult.

The baseline performance of the participants showed stronger capacity in biomedical communication tasks and comparatively weaker capacity for empathy, feedback elicitation, and confidentiality-related behaviors. This finding is consistent with prior research documenting the dominance of biomedical and paternalistic models in primary care settings, particularly in low- and middle-income countries (Jordans et al., 2022; Spedding et al., 2022). These patterns are more likely to reflect gaps in pre-service training and service structures rather than individual clinician’s skills. These findings underscore the importance of adapting competency frameworks to provider cadre, as some competencies though considered foundational may not be part of pre-service training and may require explicit instructional focus and repeated practice when introduced into primary care contexts.

The results of correlation analysis are particularly relevant for training programmes, where increased verbal activity does not necessarily translate into improved therapeutic quality. These patterns may help enhance the efficiency of training by prioritizing clusters of competencies rather than addressing skills in isolation during time-limited training.

### Limitations

Our study has some limitations and should be interpreted in light of the study’s primary aim to optimize feasibility and scalability within real-world mhGAP-HIG implementation constraints. We understand that simulated case vignettes may not reflect real-world clinical behavior or actual competence, despite being a feasible approach to assess competence. Also, video recording may introduce social desirability among physicians and may not be best reflection of their behavior in clinical settings. Additionally, use of single vignette, though efficient may not demonstrate full breath of competence/skills, and may introduce practice effect if measured pre and post training. As ENACT may show limited sensitivity to short term competency, some skills may best be assessed longitudinally.

### Future directions

Findings from the adapted ENACT assessment can inform mhGAP training design and delivery, supporting a shift from a biomedical centered to collaborative, patient-centered approach. It can guide the selection criteria for PCPs, curriculum refinement, identification of priority skills for refresher trainings, and quality improvement mechanisms for training and supervision (Jordans et al., 2022; Kohrt, Mutamba, et al., 2018). From a service delivery perspective, competency assessment can support quality of care and ethical dimensions of mental health integration into primary care by operationalizing principles such as respect, confidentiality, and collaborative engagement into observable clinical behaviors. Broader application of the adapted ENACT framework can inform policy and implementation decisions on the feasibility of integrating foundational competencies into large-scale training and supervision systems in low-resource and humanitarian contexts.

The adaptation process demonstrated in this study provides a practical template for developing culturally relevant, time-efficient assessment tools that can be sustained within routine training systems, strengthening local ownership and long-term implementation.

Future research should examine the responsiveness of the adapted ENACT tool to training-related change, its reliability and applicability to other provider cadres such as nurses or community health workers. It should also explore the impact of using the tool over a longitudinal period to assess competencies over time and the extent to which assessment-informed feedback influences routine clinical practice.

## Data Availability

The data that support the findings of this study are available from the Health Section at the Ministry of Planning, Development & Special Initiatives, Government of Pakistan. Confidentiality restrictions apply to the availability of the data used under the license for the current study, and so are not publicly available. However, the data can be made available from the authors upon reasonable request after formal approval from the Ministry of Planning, Development & Special Initiatives, Pakistan.

## Authors Contribution

AH: Conceptualization, Methodology, Visualization, Supervision, Writing – review and editing.

AsN: Methodology, formal analysis, Writing – original draft,

IH: Methodology, data collection, Writing – original draft,

AN: Methodology, data collection

NM: Data collection, writing – original draft

## Acknowledgments

we would like to thank all trainers including Dr Hazrat Ali Khan, Dr Shahnawaz Zehri, Dr. Kamal Khan Mandokhail and Dr. Hadayatullah who facilitated us in the process of implementation in Balochistan and Dr. Syed Tahir Hussain Shah for facilitating us in implementation in KP.

## Ethical Statement

This study was conducted as part of the Mental Health and Psychosocial Support Project, approved by the Ministry of Planning, Development & Special Initiatives in compliance with ethical standards and consent protocols under letter no. 6(262) HPC/2020.

## Abbreviations

ENACT: ENhancing Assessment of Common Therapeutic factors
WHO: World Health Organizations
LMICs: Low-and-middle-income countries
PCPs: Primary care physicians
mhGAP-HIG: mhGAP Humanitarian Intervention Guidelines
ICCs: Intraclass correlation coefficients

